# Cohort Profile: The China Multi-Ethnic Cohort (CMEC) Study

**DOI:** 10.1101/2020.02.14.20022970

**Authors:** Xing Zhao, Feng Hong, Jianzhong Yin, Wenge Tang, Gang Zhang, Xian Liang, Jingzhong Li, Chaoying Cui, Xiaosong Li, on behalf of the China Multi-Ethnic Cohort (CMEC) collaborative group

## Abstract

**Cohort purpose:** The China Multi-Ethnic Cohort (CMEC) is a community population-based prospective observational study aiming to address the urgent need for understanding NCD prevalence, risk factors and associated conditions in resource-constrained settings for ethnic minorities in China.

**Cohort Basics:** A total of 99 556 participants aged 30 to 79 years (Tibetan populations include those aged 18 to 30 years) from the Tibetan, Yi, Miao, Bai, Bouyei, and Dong ethnic groups in Southwest China were recruited between May 2018 and September 2019.

**Follow-up and attrition:** All surviving study participants will be invited for re-interviews every 3-5 years with concise questionnaires to review risk exposures and disease incidence. Furthermore, the vital status of study participants will be followed up through linkage with established electronic disease registries annually.

**Design and Measures:** The CMEC baseline survey collected data with an electronic questionnaire and face-to-face interviews, medical examinations and clinical laboratory tests. Furthermore, we collected biological specimens, including blood, saliva and stool, for long-term storage. In addition to the individual level data, we also collected regional level data for each investigation site.

**Collaboration and data access:** Collaborations are welcome. Please send specific ideas to corresponding author at.

## Why was the cohort set up?

Non-communicable diseases (NCDs), such as cardiovascular disease, cancer, chronic respiratory disease and diabetes, have become leading causes of death worldwide^1, 2^. It is estimated that 63% of all global deaths were due to NCDs^3^, with an increasing contribution from low- and middle-income countries. The prevalence and incidence of NCDs are not evenly distributed between and within countries because of variations in the environment, lifestyle, socioeconomic position, genetic factors, and access to health care services^4, 5^.

China has been facing a growing burden of NCDs with strong regional variation^6^. Most primary data on the prevalence of and risk factors for NCDs come from Central and Eastern China^7, 8^, with a mix of ethnicity, socioeconomic status, geography, and dietary, behavioural and lifestyle factors that is very different compared to Western China. Ethnicity in particular is increasingly seen as an important risk factor for NCDs throughout the world^9^. Ethnic minority groups are known to have different outcomes for cancer^10^, diabetes^11^, cardiovascular diseases^9^ and respiratory diseases^12^ compared to ethnic majority groups.

China, with its 55 different ethnic minority groups, represents the largest ethnic minority population in the world (114 million in 2010)^13^. However, remarkably little is known about ethnic variation in the prevalence, incidence and/or risk factors for NCDs in China. One national cross-sectional study suggested that Tibetans had a significantly lower prevalence of diabetes than the majority Han population^14^. Another national cross-sectional study found large ethnic differences in haemoglobin distribution and anaemia prevalence^15^. To our knowledge, these are the only studies examining national ethnic variation in NCDs in China.

China’s Southwest region is home to the 56 ethnic groups living in China^13^. Ethnicity not only represents genetic diversity but also characterizes strong local identities for work and daily life, such as the Tibetan people in rural areas who usually live a nomadic life. Many ethnic minorities live in remote areas, speak different dialects, and share some unique dietary habits, for example, Tibetan people favouring butter tea^16^ or the Miao people’s diet rich in sour soup (fermented acidic liquid)^17^. Although substantial efforts have been made to improve the socioeconomic status of ethnic minorities^18^, they still tend to be much poorer than their Han counterparts. Many ethnic minorities live in distinct geographical environments and climates. Tibetans, for example, live in the Tibetan Plateau with low barometric pressure, which could cause high-altitude hypoxia or decreased oxygen levels^19^. In contrast, more than 100 million Han people live in the low-altitude Sichuan Basin and are exposed to severe air pollution.

To address the urgent need for data on the prevalence of, risk factors for and associated conditions of NCDs across various ethnicities in resource-constrained settings, the China Multi-Ethnic Cohort (CMEC) study was launched. The study is based on a standardized survey approach and multiple stringent quality control (QC) measures. Such steps are essential considering not only the substantial research gap but also that the study of NCDs across multiple ethnicities is likely to reveal important and potentially novel avoidable causes, which may inform disease prevention programmes in other populations worldwide.

## Who is in the sample?

The CMEC was established in five provinces of Southwest China (Figure 1), and the baseline survey was conducted between May 2018 and September 2019. Given a full consideration of China’s ethnic characteristics, population size, and non-communicable disease patterns, we aimed to include participants from the Tibetan, Yi, Miao, Bai, Bouyei, and Dong ethnic groups. In addition, to compare those ethnic minorities to the majority of people in China, the Han ethnic population was also recruited into our cohort. A multistage, stratified cluster sampling method was used to obtain samples from community-based populations. In the first stage, one to two minority settlements for each ethnic group were selected as our study sites. To reach a better representativeness of variation with regard to geography and development state, settlements from high plateaus, basins, rural areas, and highly air-polluted regions were given special consideration. In the second stage, one to eight communities (depending on the size of communities) in each settlement were selected by the local Centres for Disease Control and Prevention (CDCs), taking into account migration status, local health conditions and, most importantly, ethnic structure. In the final stage, all participants who met our inclusion criteria were invited to participate in our studies in consideration of both sex ratio and age ratio.

**Figure 1.**
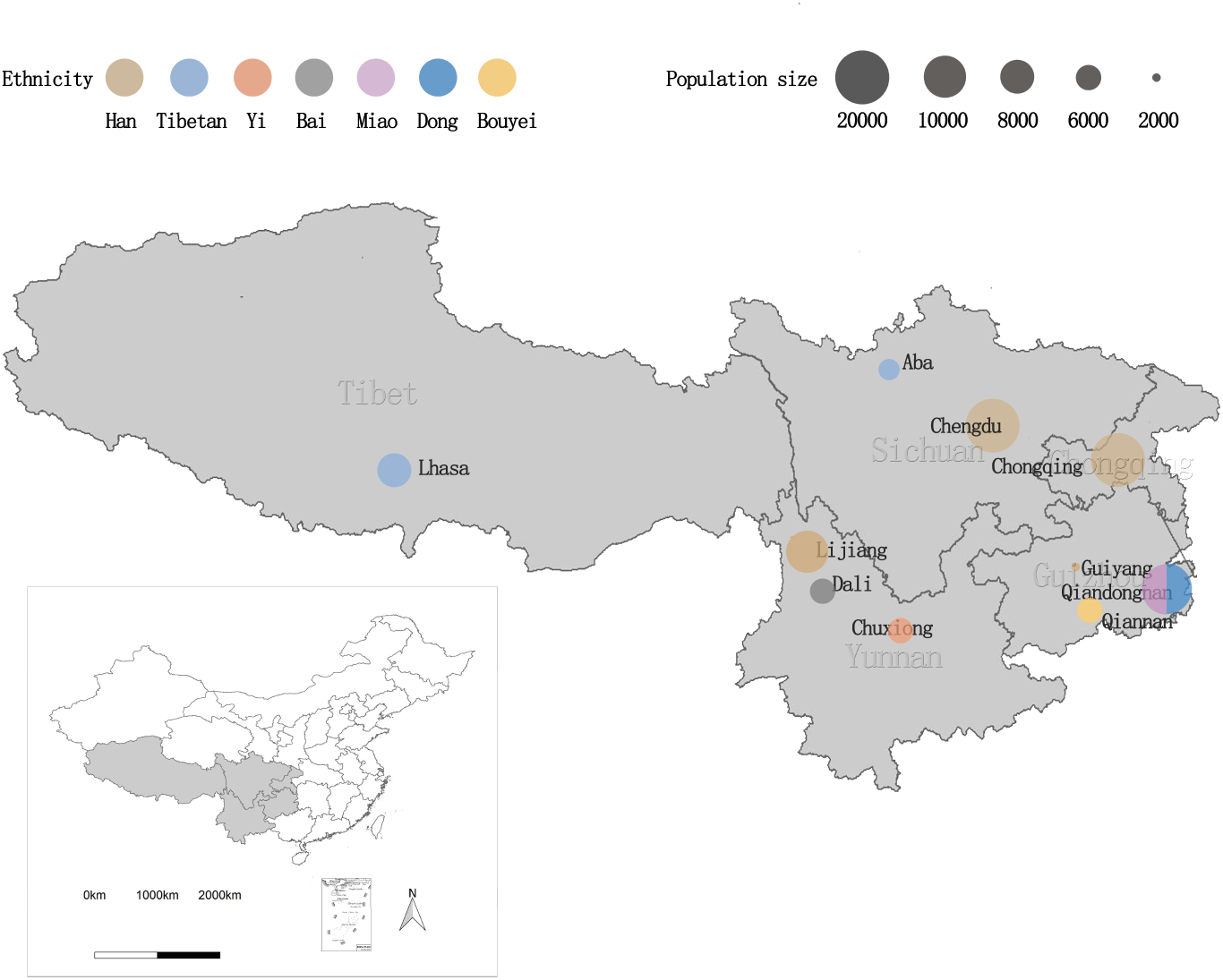
Spatial distribution of baseline populations.

The inclusion criteria we applied included (i) ages of 30-79 years on the day of the investigation (except for Tibetan populations, whose inclusion criteria for age was 18-79, given the shorter life expectancy in this population); (ii) permanent residents, capability of completing baseline surveys and availability to complete follow-up studies; and (iii) complete questionnaire interviews, physical examinations and blood tests. The exclusion criteria were as follows: (i) inability to provide a unique national identification card; (ii) severe physical or mental diseases (*e.g*., schizophrenia and bipolar disorder); and (iii) refusal to comply with the requirements of the study. All of the participants signed an informed consent form prior to data collection. Ethical approval was received from the Sichuan University Medical Ethical Review Board (K2016038).

## How often will participants be followed up, and what is the rate of loss likely to be?

The follow-up stage will be conducted in both passive and active modes. In passive mode, we will conduct follow-up via linkage to the electronic records of disease and death registries, mainly relying on the National Electronic Disease Surveillance System (NEDSS) and the National Health Insurance (HI) claim databases. Through annual data linkage with these established electronic disease registries, various health outcomes of the study participants will be obtained. The diagnoses of these conditions are based on well-accepted international standards. In China, the underlying and contributing causes of death are coded according to the 10th version of the International Statistical Classification of Diseases (ICD-10)^20^. All deaths during follow-up will be checked regularly for cause-specific mortality through the death certificates reported to the regional CDCs and the death registries. In active mode, all surviving study participants will be invited to resurvey every 3-5 years. We will re-estimate the status of risk exposure and document the incidence of multiple diseases (*e.g*., cancer, stroke, heart attack, diabetes mellitus, hypertension) using the same measurements in the baseline survey (see more details in the following section). We aim to minimize the expected rate of loss to less than an estimated 8% in the next round of follow-up.

## What has been measured?

Table 1 summarizes the measurements collected from the CMEC baseline survey, which consisted of an electronic questionnaire with face-to-face interviews, medical examinations and clinical laboratory tests. In addition, we collected biological specimens, including blood, saliva and stool, which were stored for further use. In addition to the individual level data for each participant, we also collected the regional level data for each investigation site. More details on these measurements are described below.

**Table 1.** Summary of measurements at the baseline survey in the CMEC study

| Measurements | No. of variables | Variables |
| --- | --- | --- |
| **Electronic questionnaire** |  |  |
| Demographics | 21 | National ID number, date of birth, sex, marital status, ethnic group, education, occupation, household income, household expenses, composition of household, insurance coverage, contact information, *etc.* |
| Smoking and indoor air pollution | 20 | Smoking status, smoking cessation, passive smoking exposure, pesticide exposure, indoor air pollution, *etc.* |
| Alcohol use | 11 | Frequency of drinking, types of drink, alcohol consumption, post-drinking symptoms, *etc.* |
| Tea and other beverages | 12 | Frequency of drinking tea, types of tea, frequency of drinking other beverages, types of beverage, *etc.* |
| Personal and family health | 137 | Self-rated health status (EQ-5D-5L), disease history, family disease history, defecation situation, gum situation, *etc.* |
| Physical activity | 24 | Job-related physical activity, transportation physical activity, leisure-time physical activity, housework, weight change during last 12 months, weight in early adulthood, *etc.* |
| Reproductive history (for women) | 19 | Age of menarche, menopause status, experience of pregnancy, history of breast feeding, history of contraceptive pills use, history of hysterectomy and of ovary/breast surgery, *etc.* |
| Habitual diet | 139 | Flavouring, food frequency questionnaire (FFQ) <sup>a</sup>, nutritional supplements, history of severe food |
|  |  | shortage, history of refrigerator usage, spicy food intake, Sichuan pepper intake, *etc.* |
| Life events, social support and psychological conditions | 28 | Life satisfaction, traumatic events, sleep situation, napping, probable depression (PHQ-2), probable anxiety (GAD-2), social support and social capital, *etc.* |
| **Medical examination** |  |  |
| Fasting condition | 1 | Fasting time |
| Medication history | 2 | Antihypertensive drugs, antiarrhythmic drugs |
| Anthropometric measurement | 4 | Weight, height, hip circumference, wrist circumference. |
| Physical examination | 3 | Fasting blood pressure (measured three times, OMRON HEM-8711), fasting heart rate (measured three times, OMRON HEM-8711), visual inspection (international standard visual acuity chart) |
| Lung function | 1 | Peak expiratory flow rate (KOKA PEF-3) |
| Osteopathic examination | 1 | Mineral density of anklebone (OSTEOKJ3000) |
| Electrocardiogram (ECG) | 1 | 12-lead ECG |
| Abdominal ultrasonography | 5 for male | Liver, gallbladder, spleen, pancreas, kidney |
|  | 8 for female | Uterus, ovaries and fallopian tubes (additional for female participants) |
| Chest X-ray | 2 | Cardiovascular, lungs |
| Chronic high-altitude disease <sup>b</sup> | 7 | Chronic high-altitude disease scale (7 questions or symptoms) |
| **Clinical laboratory test** |  |  |
| Routine blood | 24 | White blood cells (WBCs), red blood cells (RBCs), haemoglobin (HGB), platelets (PLTs), lymphocytes, monocytes, neutrophils, eosinophils, *etc.* |
| Fasting blood glucose | 1 | Fasting blood glucose (FBG) |
| Lipid levels | 4 | Triglycerides (TGs), cholesterol (CHOL), high-density lipoprotein cholesterol (HDL-CH) and low-density lipoprotein cholesterol (LDL-CH) |
| Hepatic function | 5 | Total protein (TP), albumin, globulin, alanine transaminase (ALT), aspartate amino transferase (AST), and gamma-glutamyl transpeptidase (GGT) |
| Kidney function | 3 | Creatinine, urea, and uric acid |
| Routine urine | 12 | Specific gravity, urobilinogen, urinary bilirubin, urinary ketone body, urine glucose, urinary protein, pH, urine nitrite, *etc.* |
| Alkaline phosphatase | 1 | Alkaline phosphatase (ALP) |
| Creatine kinase-MB | 1 | Creatine kinase-MB (CK-MB) |
| Total bilirubin | 1 | Total bilirubin (TBIL) |
| Glycosylated haemoglobin | 1 | Glycated haemoglobin (HbA1C) |
| **Biological specimens** |  |  |
| Blood sample | 6 | 2 aliquots of serum (0.5 mL × 2), 2 aliquots of plasma (0.5 mL × 2), 1 aliquot of buffy coat (0.5 mL) and 1 aliquot of blood cells (0.5 mL) |
| Saliva | 2 | 2 aliquots of saliva (1 mL × 2) |
| Stool | 1 | 1 aliquot of stool (1~2.5 g × 2) |
| **Regional level data** |  |  |
| Hourly air pollution data | 6 | Sulfur dioxide, nitrogen dioxide, nitric oxide, fine particulate matter with particle size below 10 microns (PM10), fine particulate matter with particle size below 2.5 microns (PM2.5), ozone (China National Environmental Monitoring Centre). |
| Daily meteorological data | 9 | Air pressure, air temperature, precipitation, evaporation, relative humidity, wind direction and wind speed, sunshine hours and 0 cm ground temperature elements (National Meteorological Information Centre). |
| Socioeconomic data | - | Specific variables selected according to research purposes (Yearbook data of national and provincial statistical departments) |
| Health related data | - | Specific variables selected according to research purposes (Yearbook data of national and provincial health departments) |
| Geographic data | - | Specific variables selected according to research purposes (National Earth System Science Data Sharing Platform) |

a. Includes common food items and ethnic group-specific food items.
b. Only available for high-altitude areas.

### Collaboration and field workflow

The CMEC baseline survey was conducted through the collaborative efforts of multiple agencies, including academic institutions, CDCs, local clinical centres, Third-Party Medical Laboratory (TPML) companies and local governments. Each agency has a clear role in the process of the investigation. Typically, the local government began the CMEC propaganda or publicity campaigns a few weeks before the formal investigation. Meanwhile, the residents were fully informed of the benefits and requirements of participating in the CMEC study. Then, residents who were willing to participate made an appointment with the local clinical centre. The clinic visit for each participant typically took 90–120 min. The average daily recruitment rate was 70–80 participants per site.

### Electronic questionnaire

The main content of our questionnaire referred to the study of the China Kadoorie Biobank as the prototype^21^. Major modifications were made in sections regarding physical activity, habitual diet and psychological conditions to fully capture the unique features of the different ethnic minority groups. We used a tablet computer with a self-developed application (CMEC App) to collect the questionnaire information (see more details in Table 1). The information was collected by a face-to-face interview implemented by well-trained interviewers who were typically local college students with medical backgrounds. For a skilled interviewer, an average of 30 to 45 min was often required to complete a questionnaire.

The whole interview was audio recorded, and we documented the duration spent on each question. For data QC, on the same day of data collection, the data quality inspectors, who were chosen from excellent interviewers, drew random samples of questionnaires to assess their data quality by listening to the audio records. The sampling scheme was built on our computer system and could ensure that each interviewer was sampled at least once. The next day, the assessment report was fed back to the interviewers to help them improve their interviewing skills. Questionnaires that were classified as unqualified were excluded from the final analysis.

### Medical examinations

We conducted medical examinations mainly using the resources and personnel at local clinical centres. We implemented standardized training for the doctors and nurses before the investigation.

For those testing devices that the local clinical centres were rarely equipped with, we provided unified devices for each site and trained the local staff to operate those devices, including bone mineral density densitometers (OSTEOKJ3000) and peak expiratory flowmeters (KOKA PEF-3). Particularly, for Tibet and high-altitude areas in Sichuan and Yunnan, qualified physicians used the chronic high-altitude disease scale, which consists of 7 questions or symptoms for assessing chronic high-altitude disease and its severity.

The CDC took the main responsibility of field QC guided by a handbook and serval forms to monitor key risk points. The main measures of QC included checking whether the measurement instruments were in good condition, inspecting whether the measurement was in accordance with SOPs, randomly sampling participants to re-test their results, *etc*.

### Clinical laboratory tests and biobanking

All participants provided blood and urine samples on-site at the time of the baseline survey. Venous blood samples, collected after overnight fasting (at least 8 hours), were used for clinical laboratory testing, including routine blood tests, fasting blood glucose, lipid levels and hepatic function. (Table 1). A total of 14 mL blood was collected into two ethylenediamine tetraacetic acid dipotassium (EDTA-K2) tubes (one 5 mL and one 2 mL), one EDTA-K2 tube with sodium fluoride (2 mL) and one vacuum tube without anticoagulants (5 mL). The blood samples were temporarily stored at 4°C before delivery for biobanking or testing by 2 Third-Party Medical Laboratories (TPMLs)/independent testing organizations via a cold chain. A total of 15∼20 mL mid-stream urine was collected for routine urine testing. A total of 4 000 saliva and 1 000 stool samples were collected from Tibetans and Han individuals residing in Sichuan Province for multi-omics analysis, such as the microbiome and systemic metabolism.

All the samples were centralized in a biobank located at West China School of Public Health, Sichuan University and stored at −80°C in cryogenic refrigerators. Computer systems for sample entry, access and temperature monitoring were developed and have been applied.

QC was implemented throughout the processes of sample collection, shipment, testing at the TPMLs, and storage at the biobanking facility. QC lists were strictly followed, including critical items to ensure high-quality samples, such as checking the identity of the sample and the participant ID, standardizing the volume of the samples, and regulating the storage time and temperature conditions of samples on site. To avoid any sample thawing during the shipment from each site to the biobanking facility and the TPMLs, electronic thermometers were placed in each sample’s dry ice box, and the temperature of samples was kept below −20°C before entry into the biobanking facility or testing by the TPMLs. The QC process for sample testing at the TPMLs was approved by the clinical testing centre of the China Health Committee and carried out every day during the study period. Blind panels of samples were periodically sent to the TPMLs for QC testing. Batches of controls were simultaneously put into the biobanking facility with samples and taken out for testing at regular intervals as a storage QC procedure.

### Computer system and data cleaning

All of the above measurements were stored and managed in electronic form by a self-developed computer system, with the central server located at West China School of Public Health, Sichuan University. Different datasets were linked by the unique participant’s ID. A visualized data organization schematic can be seen in Figure 2.

**Figure 2.**
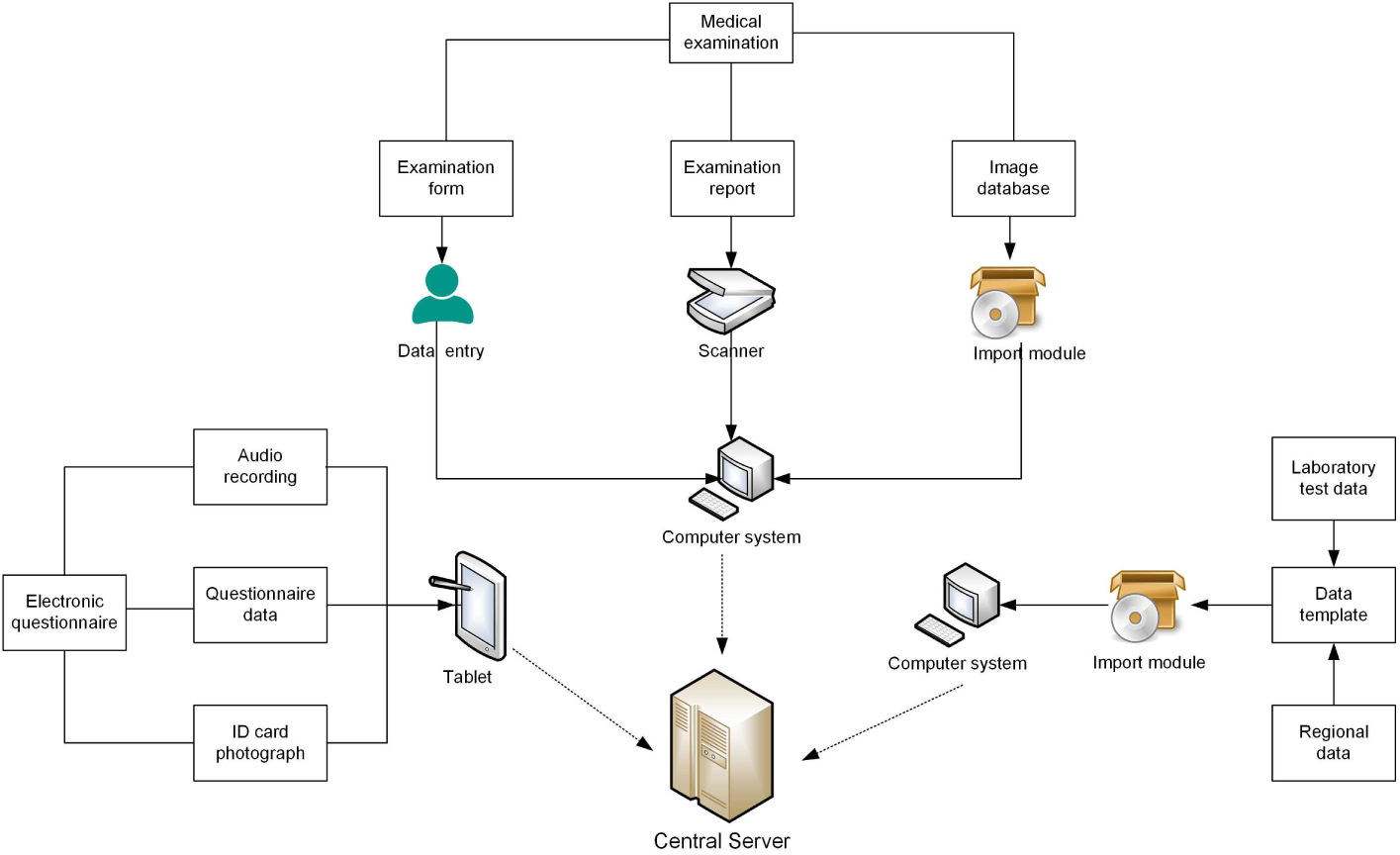
Schematic graph of the data collection and computer system.

In addition to data storage and management, our computer system also had some built-in functions to help with data cleaning based on electronic data. The whole process can be divided into two parts: detection and verification. In the detection part, we checked for duplication, completeness, outliers and logic errors via predesigned algorithms. Once suspicious data errors were detected, we ran a further verification process by listening to the audio recordings, looking up examination reports or calling back the participants. Then, the data were revised, and the whole correction process was recorded by our computer system.

## What has it found?

Table 2 shows the baseline characteristics of the participants. There were more women than men across the different ethnicities. The proportions of women ranged from 54.39% in the Han population in Basin (Chongqing, Chengdu) to 70.38% in the Bai population in the high plateau (Yunnan). While we observed a similar age distribution and marital statuses across participants of different ethnicities, significant disparities were found in terms of highest education completed. For instance, 71.40% of Tibetans in Aba reported no formal schooling, while this number was as low as 10.66% among the Han in Basin. The proportions of participants who never smoked (91.27%) or drank alcohol (93.09%) were relatively high among Tibetans in Aba compared to other populations in our study. Additionally, we noted variations regarding some lifestyle variables within an ethnic group, *e.g*., the proportion that never consumed tea was 32.31% for Tibetans in Aba and 8.12% for Tibetans in Lhasa.

**Table 2.**
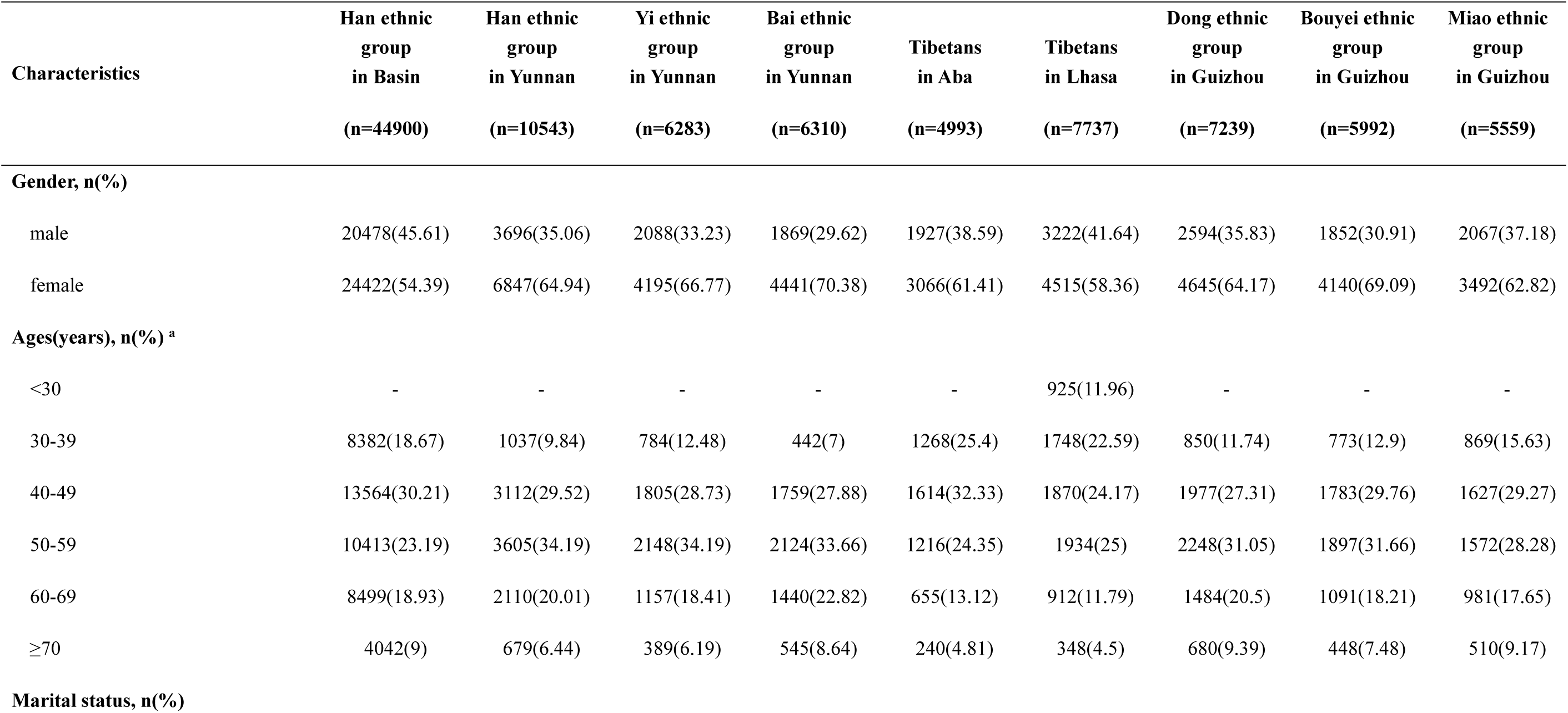
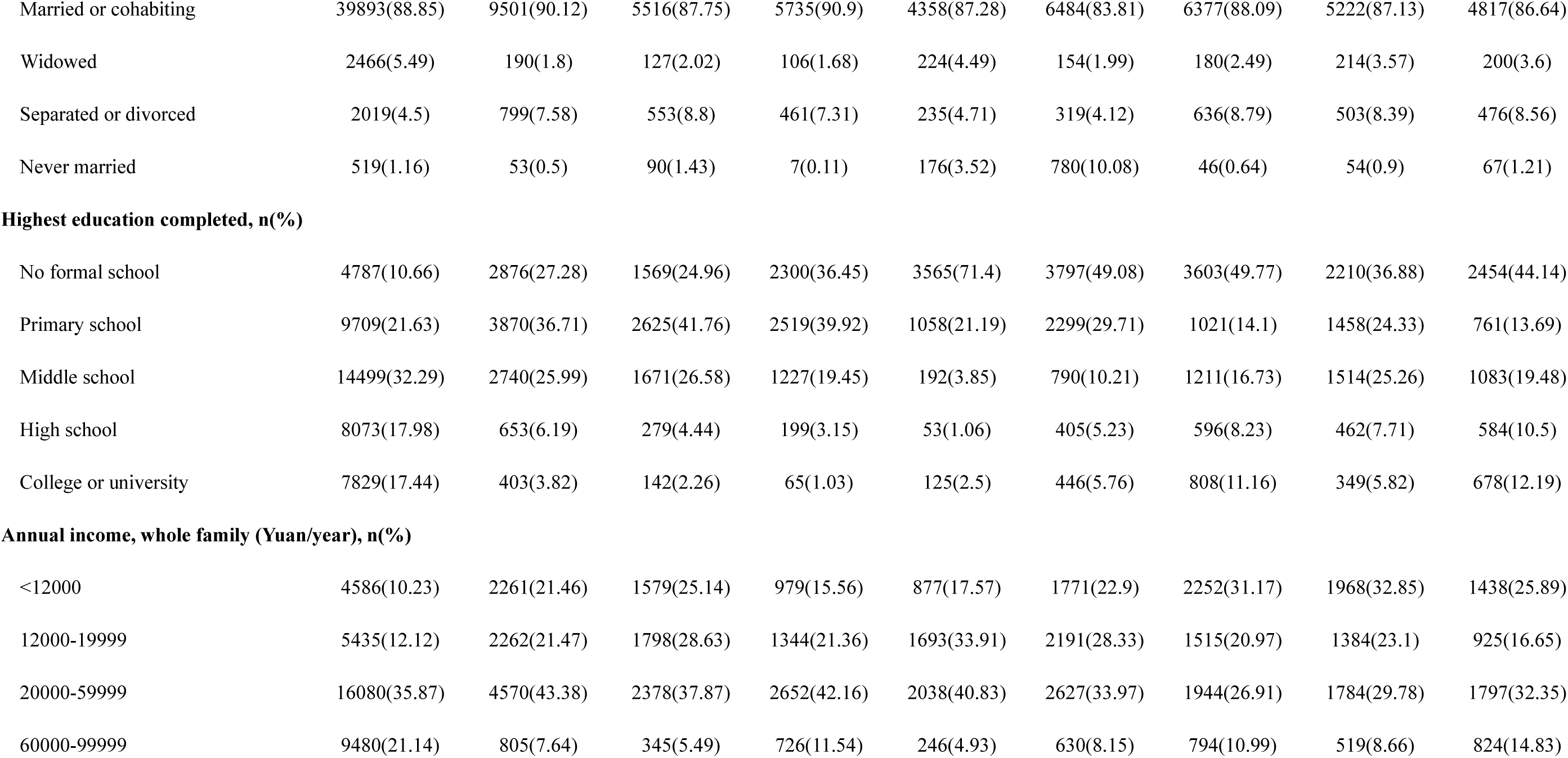
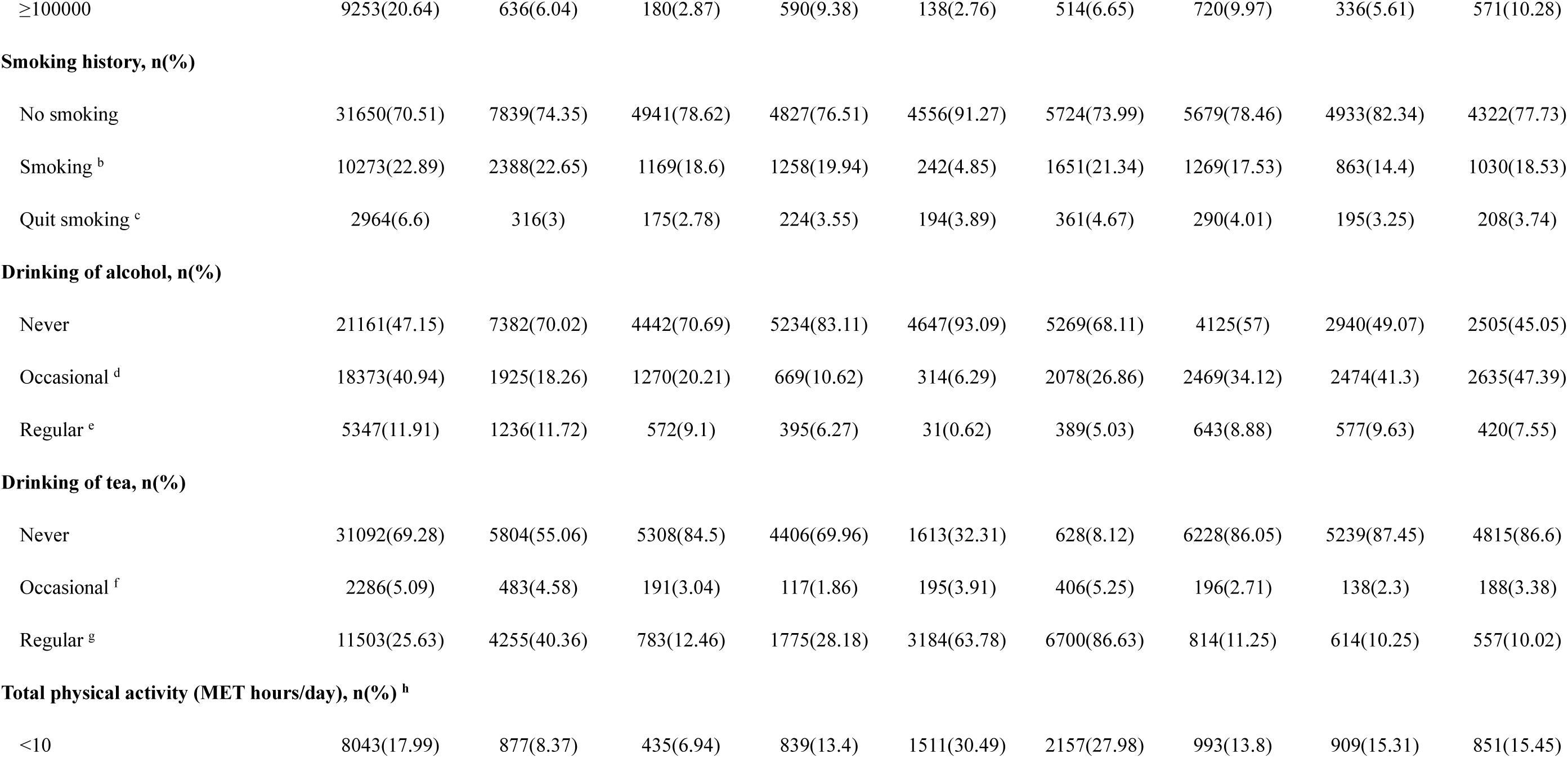
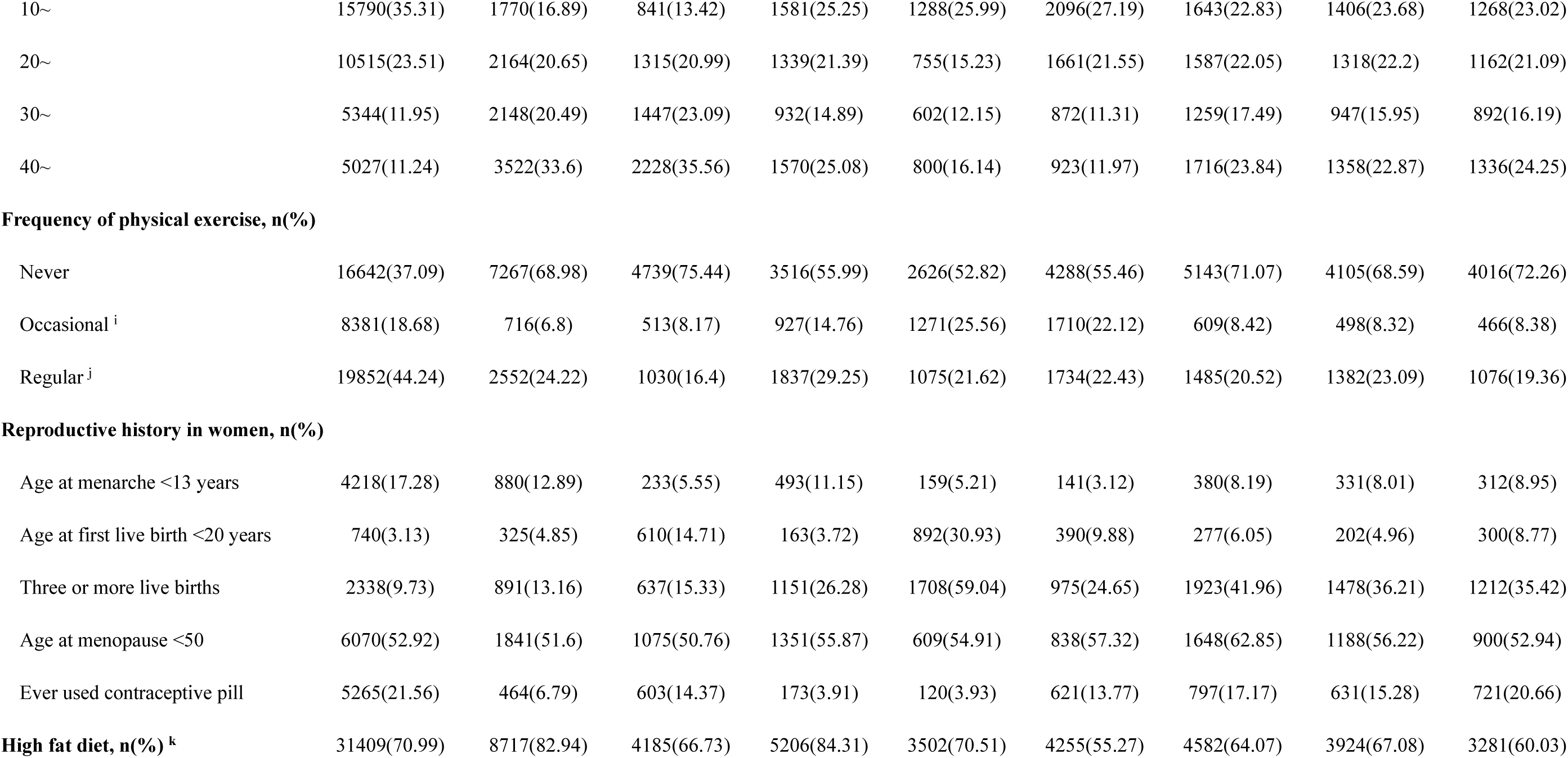
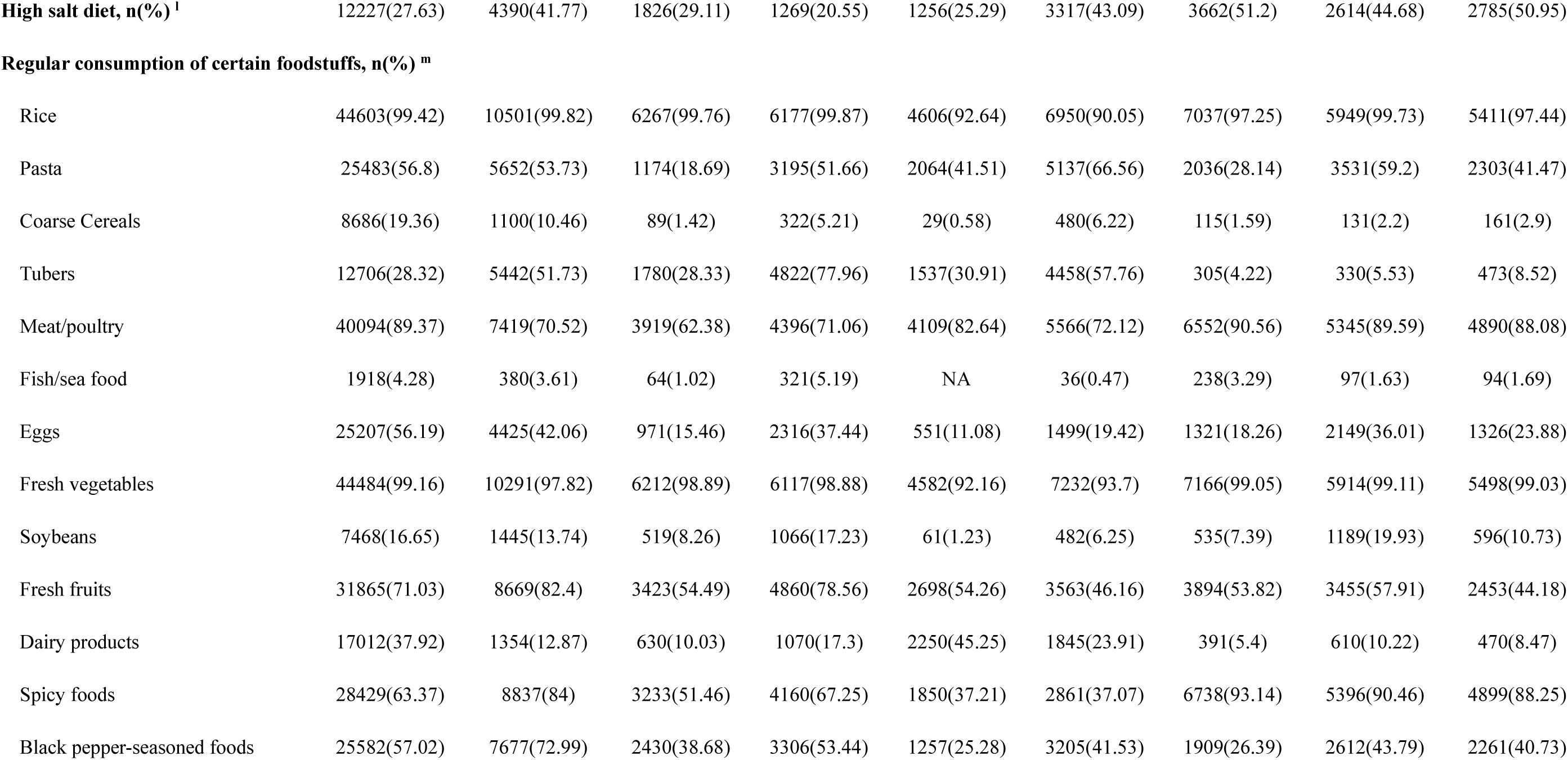
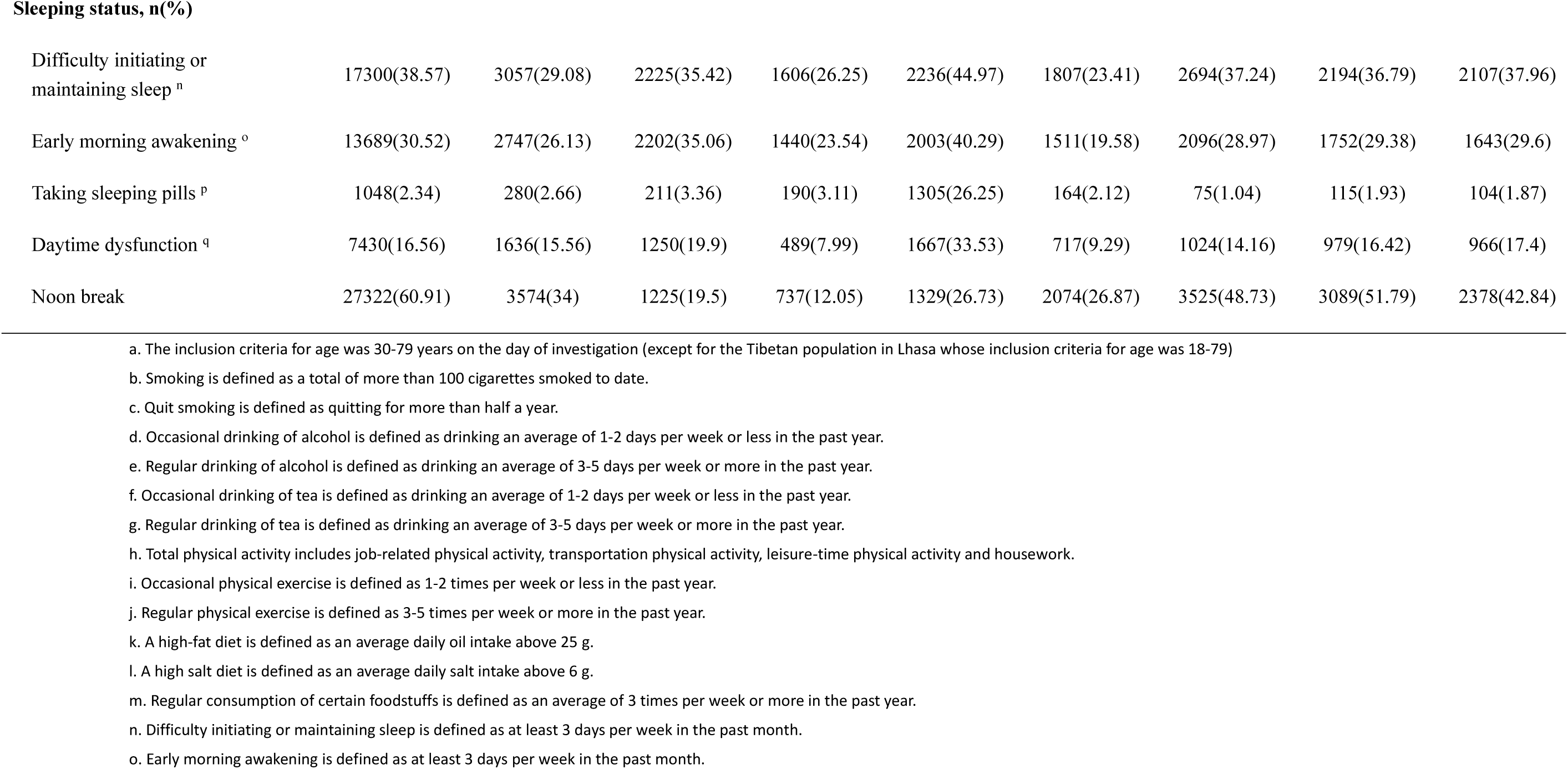
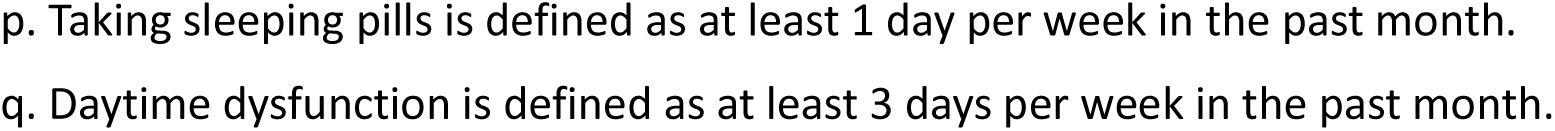
Baseline characteristics of the participants in the CMEC study

Table 3 presents the characteristics of the participants’ medical examinations. The mean height was between 154.42 cm in Miao peoples and 161.00 cm in Tibetans from Lhasa. The mean weight was between 56.93 kg in the Han population in the high plateau (Yunnan) and 65.95 kg in Tibetans from Aba. The proportion of subjects who had systolic blood pressure (SBP) higher than 140 was low (13.03%) in Lhasa Tibetans, and the proportion who had diastolic blood pressure (DBP) higher than 90 was low (15.52%) in Basin Han individuals. In terms of bone density, osteoporosis was prevalent in Aba Tibetans (14.67%) compared with other populations.

**Table 3.** Physical examinations of the participants in the CMEC study

| Characteristics | Han ethnic group in Basin | Han ethnic group in Yunnan | Yi ethnic group in Yunnan | Bai ethnic group in Yunnan | Tibetans in Aba | Tibetans in Lhasa | Dong ethnic group in Guizhou | Bouyei ethnic group in Guizhou | Miao ethnic group in Guizhou |
| --- | --- | --- | --- | --- | --- | --- | --- | --- | --- |
| Height (cm) | 158.47±8.28 | 158.25±7.54 | 157.09±7.7 | 159.27±7.7 | 158.62±8.29 | 161±8.57 | 155.35±7.77 | 155.14±7.45 | 154.42±7.94 |
| Weight (kg) | 61.67±10.47 | 56.93±9.49 | 58.26±10.64 | 58.32±9.89 | 65.95±13.44 | 65.82±10.78 | 57.81±10.3 | 58.08±10.08 | 59.65±10.11 |
| BMI (kg/m2) |  |  |  |  |  |  |  |  |  |
| < 18.5 | 832(1.89) | 777(7.67) | 351(5.71) | 437(7.1) | 60(1.25) | 116(1.86) | 310(4.66) | 218(3.92) | 110(2.15) |
| 18.5~ | 19615(44.5) | 6117(60.37) | 3262(53.1) | 3545(57.6) | 1646(34.17) | 1699(27.21) | 3230(48.53) | 2641(47.5) | 1953(38.2) |
| 24~ | 17535(39.78) | 2684(26.49) | 1846(30.05) | 1753(28.48) | 1625(33.73) | 3400(54.44) | 2339(35.15) | 2006(36.08) | 2114(41.35) |
| 28~ | 6096(13.83) | 555(5.48) | 684(11.13) | 420(6.82) | 1486(30.85) | 1030(16.49) | 776(11.66) | 695(12.5) | 935(18.29) |
| Waist circumference (cm) | 82.67±9.51 | 77.29±9.33 | 80.23±10.22 | 82.8±9.69 | 86.92±12.21 | 91.63±11.86 | 82.95±10.12 | 82.59±9.5 | 83.55±9.85 |
| Hipline (cm) | 93.75±6.69 | 89.35±6.07 | 91.88±6.65 | 92.97±6.33 | 97.58±9.14 | 100.07±8.86 | 90.96±6.34 | 92.55±6.32 | 92.23±6.39 |
| Heart rate (/min) <sup>a</sup> | 77.21±10.62 | 75.53±11.72 | 73.12±11.65 | 74.83±11.38 | 71.89±11.28 | 74.29±10.91 | 74.68±10.68 | 75.09±10.66 | 73.81±11.09 |
| SBP (mmHg) <sup>b</sup> |  |  |  |  |  |  |  |  |  |
| < 120 | 16628(37.71) | 5208(51.38) | 2126(34.6) | 2290(37.16) | 1889(39.31) | 3720(59.71) | 3009(45.25) | 2538(45.78) | 2198(43.19) |
| 120-139 | 16938(38.41) | 3308(32.64) | 2274(37.01) | 2298(37.29) | 2045(42.55) | 1698(27.26) | 2282(34.32) | 1778(32.07) | 1763(34.64) |
| ≥140 | 10533(23.88) | 1620(15.98) | 1745(28.4) | 1574(25.54) | 872(18.14) | 812(13.03) | 1358(20.42) | 1228(22.15) | 1128(22.17) |
| **DBP (mmHg) <sup>c</sup>** |  |  |  |  |  |  |  |  |  |
| < 80 | 24697(56.01) | 5671(55.94) | 3149(51.25) | 3414(55.4) | 2726(56.8) | 3464(55.65) | 3476(52.25) | 2579(46.45) | 2450(48.11) |
| 80-89 | 12555(28.47) | 2771(27.34) | 1719(27.98) | 1721(27.93) | 1277(26.61) | 1637(26.3) | 1894(28.47) | 1720(30.98) | 1497(29.39) |
| ≥90 | 6843(15.52) | 1695(16.72) | 1276(20.77) | 1027(16.67) | 796(16.59) | 1124(18.06) | 1283(19.28) | 1253(22.57) | 1146(22.5) |
| **Pulmonary**<br>**function (L/min)** | 355.76±125.1 | 349.52±100.58 | 328.88±104.29 | 286.62±89.07 | 292.35±132.59 | 200.63±94.81 | 360.96±94.25 | 348.94±84.75 | 366.42±97.03 |
| **Bone density** |  |  |  |  |  |  |  |  |  |
| Normal | 23976(54.38) | 6421(63.32) | 4330(70.45) | 3331(54.06) | 2922(60.74) | 4043(64.84) | 3995(59.98) | 3503(62.85) | 3035(59.3) |
| Osteopenia | 17082(38.74) | 3479(34.31) | 1683(27.38) | 2490(40.41) | 1183(24.59) | 1686(27.04) | 2435(36.56) | 1844(33.08) | 1893(36.99) |
| Osteoporosis | 3034(6.88) | 240(2.37) | 133(2.16) | 341(5.53) | 706(14.67) | 506(8.12) | 231(3.47) | 227(4.07) | 190(3.71) |

a, b, c. Heart rate and systolic and diastolic blood pressure (SBP and DBP) were measured three times for each participant, and the statistics are based on the average of the three measurements.

Table 4 indicates the biochemical characteristics of the participants measured at baseline. The mean fasting blood glucose (FBG) was significantly low in Tibetans from Lhasa (4.53 mmol/L) and Tibetans from Aba (4.62 mmol/L). The mean HbA1C was between 5.69% and 6.01% across different ethnicities. With regard to triglycerides (TG), the highest TG (1.97 mmol/L) was measured among the Han ethnic group in the high plateau (Yunnan), while the lowest TG (1.00 mmol/L) was found among Tibetans in the high plateau of Aba.

**Table 4.** Biochemical characteristics of the participants in the CMEC study

| Biochemical indexes | Han ethnic group in Basin | Han ethnic group in Yunnan | Yi ethnic group in Yunnan | Bai ethnic group in Yunnan | Tibetans in Aba | Tibetans in Lhasa | Dong ethnic group in Guizhou | Bouyei ethnic group in Guizhou | Miao ethnic group in Guizhou |
| --- | --- | --- | --- | --- | --- | --- | --- | --- | --- |
| FBG (mmol/l) | 5.51±1.49 | 5.15±1.24 | 5.56±1.37 | 5.46±1.2 | 4.62±1.1 | 4.53±1.31 | 5.55±1.36 | 5.42±1.37 | 5.5±1.35 |
| ALP (U/l) | 86.66±29.39 | 84.08±27.73 | 83.99±29.01 | 90.24±28.48 | 114.71±43.52 | 113.24±37.92 | 92.56±32.68 | 74.27±23.77 | 81.19±26.88 |
| TBIL (μmol/l) | 12.65±7.1 | 11.45±5.72 | 10.17±5.05 | 12.03±6.33 | 12.73±8.5 | 13.86±6.9 | 9.92±5.11 | 10.07±4.56 | 10.27±5.36 |
| HBA1C (%) | 5.71±0.94 | 5.83±0.84 | 5.74±0.76 | 5.76±0.77 | 5.93±0.83 | 6.01±1.04 | 5.74±0.97 | 5.79±0.97 | 5.69±0.95 |
| WBC (10<sup>9</sup>/l) | 5.87±1.67 | 6.64±2.04 | 6.41±1.75 | 6.43±1.68 | 5.73±1.58 | 6.18±1.72 | 6.56±1.77 | 6.15±1.65 | 6.49±1.8 |
| RBC (10<sup>12</sup>/l) | 4.78±0.55 | 4.97±0.48 | 4.8±0.47 | 5.02±0.49 | 5.32±0.59 | 5.08±0.81 | 4.73±0.59 | 4.7±0.59 | 4.8±0.56 |
| HGB (g/l) | 144.19±16.44 | 150.63±15.12 | 147.8±14.72 | 151.55±15.46 | 160.29±21.22 | 154.45±28.46 | 140.75±16.45 | 137.35±16.21 | 143.61±16.07 |
| PLT (10<sup>9</sup>/l) | 192.88±67.78 | 217.14±59.4 | 232.58±63.08 | 203.41±62.05 | 240.14±64.6 | 233.27±88.28 | 217.59±71.26 | 227±70.9 | 239.17±65.94 |
| TG (mmol/l) | 1.64±1.42 | 1.97±1.74 | 1.89±2.09 | 1.56±1.26 | 1±0.58 | 1.27±0.83 | 1.95±1.79 | 1.76±1.6 | 1.9±1.86 |
| CHOL (mmol/l) | 4.98±1 | 5.02±0.98 | 5.51±1.14 | 5.11±0.97 | 4.82±0.97 | 4.65±0.99 | 4.94±0.99 | 4.96±0.96 | 5.02±0.98 |
| HDL-CH (mmol/l) | 1.45±0.39 | 1.48±0.4 | 1.57±0.44 | 1.69±0.44 | 1.32±0.28 | 1.29±0.28 | 1.49±0.39 | 1.53±0.29 | 1.48±0.34 |
| LDL-CH (mmol/l) | 2.88±0.78 | 2.98±0.83 | 3.52±1 | 3.04±0.84 | 3.13±0.81 | 2.8±0.74 | 2.98±0.88 | 2.51±0.7 | 2.88±0.85 |
| **TP (g/l)** | 76.26±4.55 | 75.56±4.64 | 76.22±4.75 | 73.98±4.34 | 77.58±5.07 | 76.54±5.31 | 79.22±4.62 | 77.51±4.53 | 78.58±5.36 |
| **ALT (U/l)** | 24.24±19.03 | 20.51±16.49 | 21.41±15.99 | 23.29±15.78 | 30.76±47.71 | 32.3±26.18 | 24.9±21.56 | 24.52±20.41 | 23.9±19.35 |
| **AST (U/l)** | 26.13±13.54 | 25.6±12.46 | 25.38±14.58 | 28±13.16 | 28.44±24.51 | 29.35±18.18 | 28.58±18.45 | 27.97±17.66 | 27.68±15.56 |
| **GGT (U/l)** | 34.21±49.62 | 39.7±63.11 | 47.2±92.03 | 38.78±54.15 | 42.28±46.62 | 52.38±90.48 | 43.85±80.96 | 38.75±53.1 | 41.07±68.46 |
| **Cr (μmol/l)** | 67.92±18.44 | 78.3±20.37 | 76.56±20.77 | 72.07±18.9 | 65.81±16.73 | 66.57±15.04 | 70.42±29.67 | 64.31±24.32 | 66.75±24.75 |
| **UREA (mmol/l)** | 5.46±1.51 | 5.06±1.48 | 5.26±1.57 | 5.16±1.5 | 5.2±1.5 | 4.7±1.38 | 5.18±1.67 | 5.29±1.64 | 5.08±1.66 |
| **UA (μmol/l)** | 322.25±86.65 | 318.55±87.74 | 299.82±87.69 | 286.09±83.08 | 316.72±89.8 | 334.31±89.25 | 337.6±96.57 | 315.7±91.51 | 336±95.32 |
| **CK-MB (U/l)** | 16.66±13.75 | - | - | - | 16.69±9.05 | - | - | - | - |

Figure 3 presents the prevalence of self-reported diseases, which demonstrates the diverse disease patterns among different ethnic groups. The highest prevalence of hypertension was observed in the Bai (20.50%) and Yi (20.17%) ethnic groups in the high plateau, while the lowest was reported in Tibetans from Aba (11.41%). Diabetes was mostly prevalent among the Han ethnic group in Basin (6.52%). Regarding coronary heart disease, substantial disparity was observed. The most prevalent group was Tibetans from Lhasa (3.69%), while the least prevalent group was their rural counterpart, Tibetans from Aba (0.16%). Overall, cancer was not frequently reported, with the highest observation in the Bouyei ethnic group in Guizhou (1.37%).

**Figure 3.**
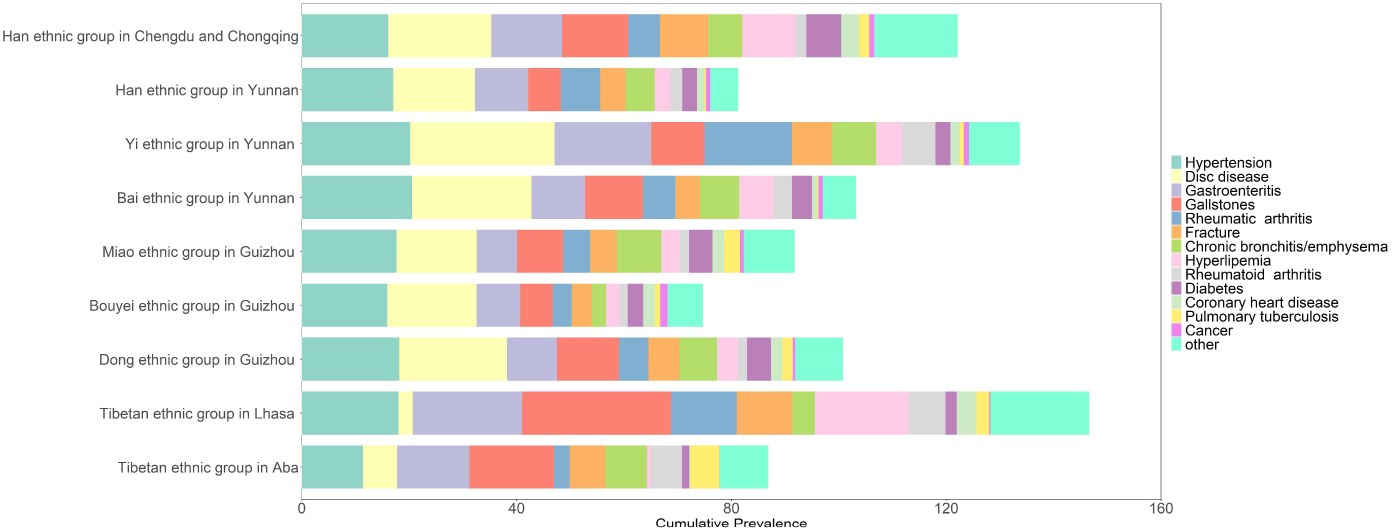
Prevalence of some selected self-reported diseases among the participants in different ethnic groups.

## What are the main strengths and weaknesses?

This study has some limitations worth mentioning. First, we relied on self-reported data for some variables, such as lifestyle factors, which may incur recall bias. Second, we excluded populations aged outside of 30-79 (except for the Tibetan populations in which 18-30-year-olds were also included), which may have ignored information on early life exposures. Despite the above limitations, our study is unique. First, to our knowledge, this is the first large-scale cohort study focusing on ethnic minority groups in China. Due to the substantial disparity in terms of social and economic development among the ethnic minority groups, as well as their distinct and diverse exposures and outcomes, the findings based on our cohort study have the potential to not only add evidence to the extant body of literature on the prevalence and risk factors of certain NCDs but also reveal novel knowledge that may serve as a reference to other global populations. Second, our study involved multi-ethnic individuals, involving mixed groups of urban and rural residents, plateau and basin inhabitants, populations living in highly concentrated and air polluted areas and those in remote mountainous areas. This would allow one to conduct comparative studies of the effects of various environmental exposures on people’s health outcomes from different angles. For instance, Tibetan populations in rural areas (Aba prefecture) live a nomadic life, which is not shared by their urban counterparts (those in Lhasa). One thus will be able to investigate significant disparities of lifestyle even within the same ethnic group. Third, our baseline survey collected detailed and comprehensive information. For instance, in addition to demographics, socioeconomics, lifestyles, and health-related histories, we also collected biological specimens and performed medical examinations and clinical laboratory tests for every participant. Fourth, our study is funded by the National Key Research and Development Program. As the highest level research program in China, this study is strongly supported by the different levels of government, CDCs and local clinic centres. Therefore, related resources, field surveys, public cooperation and electronic medical records checks can be guaranteed and thus may improve the data quality and minimize the loss of participants at follow-up. Fifth, follow-up information will be obtained through annual data linkage to electronic records of disease and death registries, as well as re-interviews every 3-5 years. This follow-up strategy enables accurate and reliable tracking of both exposures and outcomes. Finally, various methods have been rigorously implemented at every step to ensure the quality of the data.

## Can I get hold of the data? Where can I find out more?

To maximize the use of the CMEC data, we welcome collaboration from all over the world. Currently, the database is not accessible to be downloaded publicly because of some sensitive information. However, researchers interested in our study could contact the corresponding author at by providing specific ideas for more information.

**Profile in a nut shell**

- The China Multi-Ethnic Cohort (CMEC) is a community population-based prospective observational study aiming to address the urgent need for understanding NCD prevalence, risk factors and associated conditions in resource-constrained settings for ethnic minorities in China.
- A total of 99 556 participants aged 30 to 79 years (Tibetan populations include those aged 18 to 30 years) from the Tibetan, Yi, Miao, Bai, Bouyei, and Dong ethnic groups in Southwest China were recruited between May 2018 and September 2019.
- All surviving study participants will be invited for re-interviews every 3-5 years with concise questionnaires to review risk exposures and disease incidence. Furthermore, the vital status of study participants will be followed up through linkage with established electronic disease registries annually.
- The CMEC baseline survey collected data with an electronic questionnaire and face-to-face interviews, medical examinations and clinical laboratory tests. Furthermore, we collected biological specimens, including blood, saliva and stool, for long-term storage. In addition to the individual level data, we also collected regional level data for each investigation site.
- Collaborations are welcome. Please send specific ideas to corresponding author at.

## Data Availability

Currently, the database is not accessible to be downloaded publicly because of some sensitive information. However, researchers interested in our study could contact the corresponding author at by providing specific ideas for more information.

## Acknowledgements

The most important acknowledgement is to all the participants in the study, as well as to all the interviewers and members of the survey teams in each of the study sites and communities. We also thank Drs Carine Ronsmans, Peng Jia and Huan Song for their critical reading of the manuscript.

